# Ambulosono-Enhanced Ankle Rotation Training Leads to Significant Gains in Balance Among Healthy Adults

**DOI:** 10.1101/2024.04.30.24306658

**Authors:** Bin Hu, Muhammad Farhan Raza, Dhruvil Patel, Shahryar Wasif, Taylor Chomiak

## Abstract

The ankle joint, a pivotal element in lower limb-ground interactions, plays a critical role in maintaining gait and balance. In this study, we utilized the Ambulosono device—a sensor-based, music-contingent digital tool designed to assist and monitor ankle training—to investigate the effects of ankle rotation training on functional balance. We measured the durations of the single-leg stand test (SLST) under eyes-closed conditions in a cohort of healthy young adults. Comparisons of pre- and post-training SLST durations were made between the trained and untrained legs within the same subjects. Our findings demonstrated a substantial increase in the SLST durations ipsilateral to the trained ankles, while the untrained ankles in the control legs showed no significant changes. This enhancement in balance function was observed to persist for several hours post-training.

## Introduction

The ankle joint is a hinged synovial joint formed by the juncture of the talus, tibia, and fibula bones creating the kinetic connection that enables the lower limb to interact with the ground—a crucial component of balance.1, 2 Two movements of the ankle that occur at the tibiotalar joint in the sagittal plane are dorsiflexion and plantarflexion. Dorsiflexion is the movement that involves bending the foot towards the shin, thus decreasing the angle between them.1, 3 In contrast, plantarflexion refers to moving the foot downwards by lifting the heel of the foot from the ground.1 Importantly, supine ankle rotation training is a targeted exercise that strengthens the muscle groups responsible for both dorsiflexion and plantarflexion, enhancing the joint’s overall functionality.4

Maintaining balance during the Single Leg Stand Test (SLST) involves a complex interplay of sensory, motor, and cognitive functions.5 The test requires the individual to stand on one leg while keeping the other foot off the ground, challenging the body’s proprioceptive system to maintain a stable posture.5 During SLST, the ankle joint plays a pivotal role as it constantly adjusts to minor shifts in body weight and position. These adjustments are facilitated by the ankle’s dorsiflexor and plantarflexor muscles, which work in tandem to stabilize the joint. The ability to successfully perform the SLST is not only indicative of good ankle joint functionality but also reflects the efficiency of the neuromuscular system in coordinating balance.5

Poor balance can profoundly affect an individual’s physical health, functional capabilities, and overall well-being. Research underscores several potential consequences of poor balance, such as the following. First, individuals with poor balance face a heightened risk of falls, leading to severe injuries such as fractures, head trauma, and soft tissue damage.6, 7 This risk is especially acute for older adults, for whom falls can result in a loss of independence, diminished quality of life, and escalating healthcare costs.8,9 Second, poor balance can adversely affect one’s ability to carry out daily tasks, like walking, climbing stairs, or rising from a chair, thereby limiting functional mobility.10,11 Third, those with poor balance may find it challenging to engage in physical activities and sports, impacting both their overall fitness and social interaction.12,13 Additionally, poor balance has been linked to a decrease in self-confidence and an increased fear of falling, which can lead to a sedentary lifestyle.14 Lastly, studies indicate that poor balance correlates with cognitive impairments, affecting attention, memory, and executive function.15, 16 In summary, the ramifications of poor balance go beyond mere physical limitations, significantly affecting an individual’s overall health and well-being Emerging evidence suggests that ankle range of motion (ROM) is a critical factor in maintaining balance and preventing further injuries, especially among older adults.17, 18 Limited ankle ROM has been associated with an increased risk of balance-related challenges, including instability during daily movements and activities. Evaluating lower extremity ROM is therefore integral for assessing joint health and its impact on an individual’s ability to maintain balance, which is of particular concern for the elderly.19

This measure has also been deemed a useful predictor for the risk of lower limb injuries.20,21, 22 Consequently, the development of effective exercise methods targeting ankle ROM and balance is of great importance, especially for older adults concerned with preventing falls and injuries.

The Ambulosono device provides a sensor-based method for measuring and training ankle ROM.23 Understanding whether using the Ambulosono for ankle training can improve SLST performance has implications for rehab, physiotherapy, and overall mobility enhancement.

Given the link between ankle ROM and SLST performance, there is a need to evaluate the impact of Ambulosono training on both ankle ROM and SLST results. By doing so, we may unlock new strategies for balance training, injury prevention, and mobility enhancement.

In this study, we investigate the direct impact of targeted ankle joint rotation training on balance, as measured by the Single Leg Stand Test (SLST) performance, in a cohort of healthy young adults. Employing the Ambulosono device, a sensor-based tool designed to facilitate and monitor ankle training, this research aims to provide a quantitative analysis of the training’s effectiveness. By measuring pre- and post-training SLST durations, the study seeks to evaluate the specific contribution of enhanced ankle ROM to balance improvement. Additionally, the study utilizes advanced predictive modeling techniques to analyze real-world data from participants and extends its analysis through the generation and examination of a large synthetic dataset, enabling a comprehensive exploration of factors influencing balance.

## Methods

Our study was carried out via Open Digital Health (OpenDH) program, a student research training initiative aimed at providing early exposure to learning and applying advanced research and data analytics skills. Our overarching strategy centers on student education through the integration of advanced data analytics and statistical training, leveraging the capabilities of Generative Pre-trained Transformer (GPT) models.

The scalability of GPT-based training, such as ChatGPT-4, can significantly enhance accessibility to advanced statistics education. Students from diverse backgrounds and disciplines can access high-quality, interactive training resources online, breaking down barriers to advanced education in statistics and research. By providing an interactive, personalized learning environment, OpenDH program fosters a deeper understanding of statistical concepts among students, while simultaneously exposing them to cutting-edge AI tools and methodologies.

### Participants

Eighteen healthy individuals, aged 16 to 24 years, were recruited for this study. All subjects were free from injury and had no history of hospitalization or chronic disease influencing their ankle rotation and balance capacity. Moreover, they were not involved in any competitive sport and they were lifetime non-smokers. Ethics approval was obtained from the University of Calgary Research Ethics Board as part of Ambulosono registered trial (ISRCTN06023392). Informed written consent was obtained from participants at baseline prior to participation.

### Pre-training SLST Assessment

During the baseline assessment of SLST the participants stood on one leg with their eyes closed. The test began as soon as the non-weight-bearing leg was lifted off the ground and ended if the participant opened their eyes, touched the ground with the non-weight-bearing leg, or if the weight-bearing leg lost contact with the ground. Each leg was tested once, with an additional attempt allowed for durations less than 2 seconds. The leg with the shorter balance time was identified as the weak, lower-performing leg targeted for training.

### Ambulosono Device and Data Capture

Ambulosono is a single sensor-based motion tracking device consisting of 3-axis MEMS-based gyroscopes and 3 accelerometers with validated accuracy (Chomiak et al. 2019).

Ambulosono App uses fusion codes for automatic gravity calibrations and real-time angle output (pitch, roll, and yaw) with high accuracy. Motion data was automatically captured by using circumference geometry and radius and radians to functionally relate ankle flexion in real-time at a sampling frequency of 50 Hz [7]. The App can also detect the amplitudes of joint motions and use the signals to switch “on” or “off” different playlists of highly motivational music. Therefore, users can use self-generated music cues as feedback signals to maintain a pre-defined, amplitudes of ROM. Training data is automatically archived in the phone and transmitted via Wi-Fi to a secured server with level 3 encryption.

### Training Protocol

Prior to training, the subjects were taught on how to use Ambulosono device to record dorsiflexion and plantarflexion and how to use Ambulosono music feedback to achieve and maintain a minimal range of ankle rotations. Participants were instructed to perform maximum ankle dorsiflexion and plantar extension, either in a supine or sitting position, with the Ambulosono sensor wrapped around the dorsum of the foot of the weaker leg that has a shorter SLST. The stronger leg, which was not engaged in ankle training, served as a within subjects’ control. Each participant completed 90 minutes of training at home at their own schedule over a three-day period. The daily training sessions, duration, and “dosage” were recorded by Ambulosono device for further analysis.

### Post-Training SLST Assessment

Two post-training SLST were conducted using the same protocol as described for the baseline assessment. They were administered at two separate time points within the five days after participants completed the 90 min training. For remote testing sessions, SLST assessments were conducted via online video calls, ensuring full body visibility of participants. Two administrators managed these sessions: one timing the test and the other ensuring adherence to the protocols. The time stamps of the post-training SLST were used to estimate the wash-out effect of ankle training.

### Data Analysis

In this study, we used ChatGPT and its Advanced Data Analysis Platform in several aspects of the result analysis. First, a specific module of data integrity control was created to detect and identify errors related to missing data, duplicates and discrepancies, which can occur during manual handling and cleaning of research data (e.g. during cut and paste among different spreadsheets). Our data integrity module, which was created using python scripts based on natural language instructions, automatically checks and validates the accuracy of numerical data and timestamps among multiple data sheets including the intermediate files created during statistical analysis. Second, given the unique strength LLMs in translating natural language into statistical coding, were able to perform cross-validations using different statistical inference (e.g. regression and mixed-effect models) and machine learning models. Finally, GPT platform also allowed us to create synthetic databases based on real world data for modeling and simulation. ChatGPT’s role underscores the evolving landscape of AI in academic research, demonstrating how such tools can significantly contribute to complex scientific endeavors.

## Results

The flowchart progresses through the initial baseline assessment to identify the lower-performing leg, followed by 90 minutes Ambulosono training protocol over three days with self-selected schedule. Post-training assessment was conducted within five days after training.

**Figure 1.**
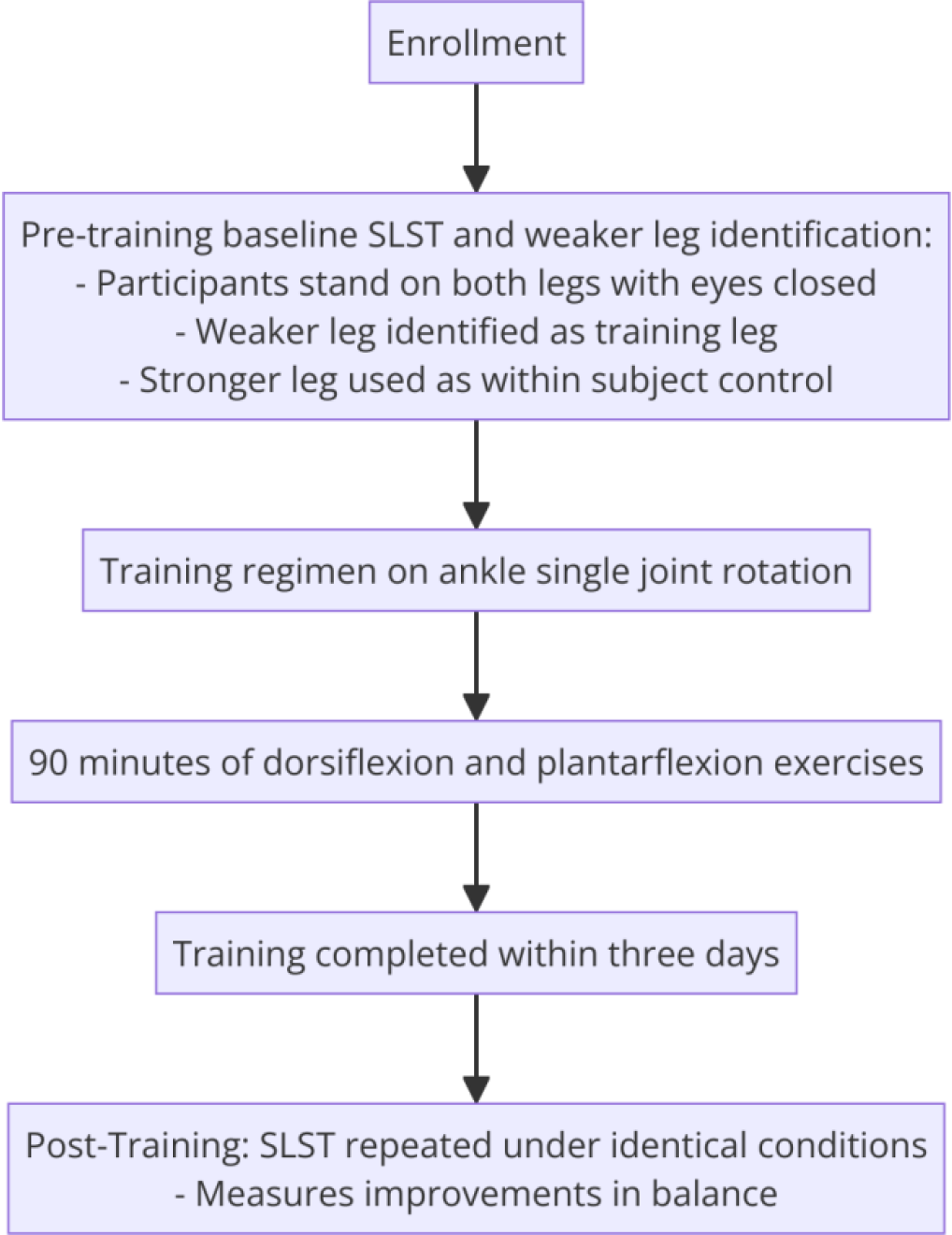
Study Protocol illustrating baseline SLST tests, training leg identification, and the regimen of ankle rotation training for all participants.

**Table 1:** This table illustrates the demographic categories, including gender, age group, height, and leg length. Percentages indicate the proportion of participants within each category.

| Demographic Category | Percentage |
| --- | --- |
| Total Participants | 100% |
| Gender: Male | 47.37% |
| Gender: Female | 52.63% |
| Age Group: <18 | 9.09% |
| Age Group: 18-20 | 63.64% |
| Age Group: >20 | 27.27% |
| Height: Short (<160 cm) | 27.27% |
| Height: Average (160-170 cm) | 9.09% |
| Height: Tall (170-180 cm) | 27.27% |
| Height: Very Tall (>180 cm) | 36.36% |
| Leg Length: Short (<0.75) | 0.00% |
| Leg Length: Average (0.75-0.85) | 18.18% |
| Leg Length: Average (0.85-0.95) | 18.18% |
| Leg Length: Long (0.95-1.05) | 63.64% |
| Leg Length: Very Long (>1.05) | 0.00% |

In this study, each of the 18 participants first underwent a baseline SLST assessment, with the lower-performing leg with shorter SLST duration identified as the training leg. Our training regimen consist of 90 minutes of continued ankle dorsiflexion and plantarflexion to be completed within a three-day period.

As shown in Figure 2, ankle rotation training resulted in a substantial increase in SLST that was only detectable in the training leg. Sign Test indicated a significant increase (p <0.001) with a with a large effect size (Cliff’s Delta=0.9883)

**Figure 2.**
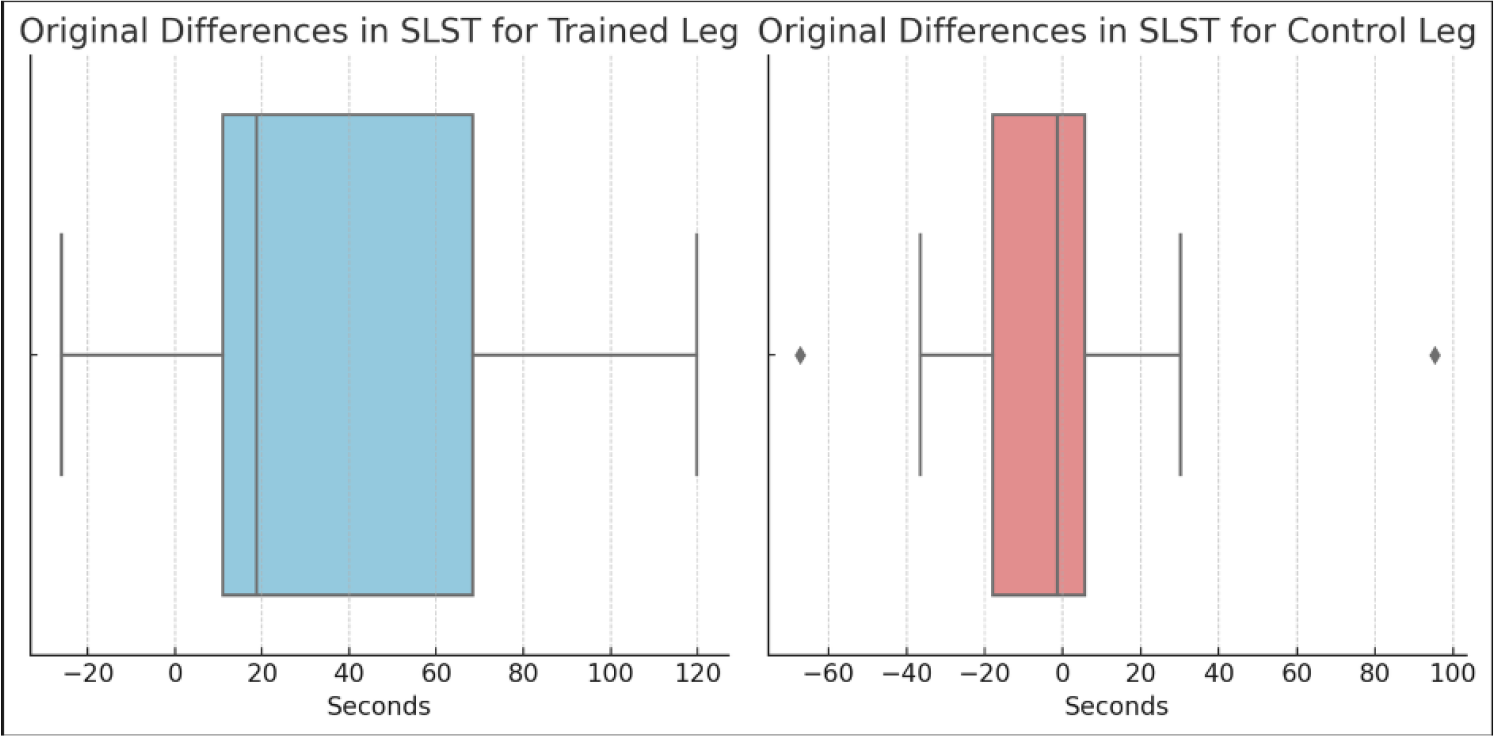
Comparison of SLST duration improvements between trained and control legs. Box plots show the distribution of differences in SLST scores before and after the training regimen. The left box plot (blue) shows changes for the trained legs, where a positive shift indicates improved balance performance post-training. The right box plot (red) illustrates changes for the control legs, with the median line close to zero reflecting no significant change in balance performance. Outliers are represented by diamond shapes. The greater improvement in trained legs compared to control legs highlights the effectiveness of the ankle single joint rotation exercises in enhancing balance.

**Figure 3.**
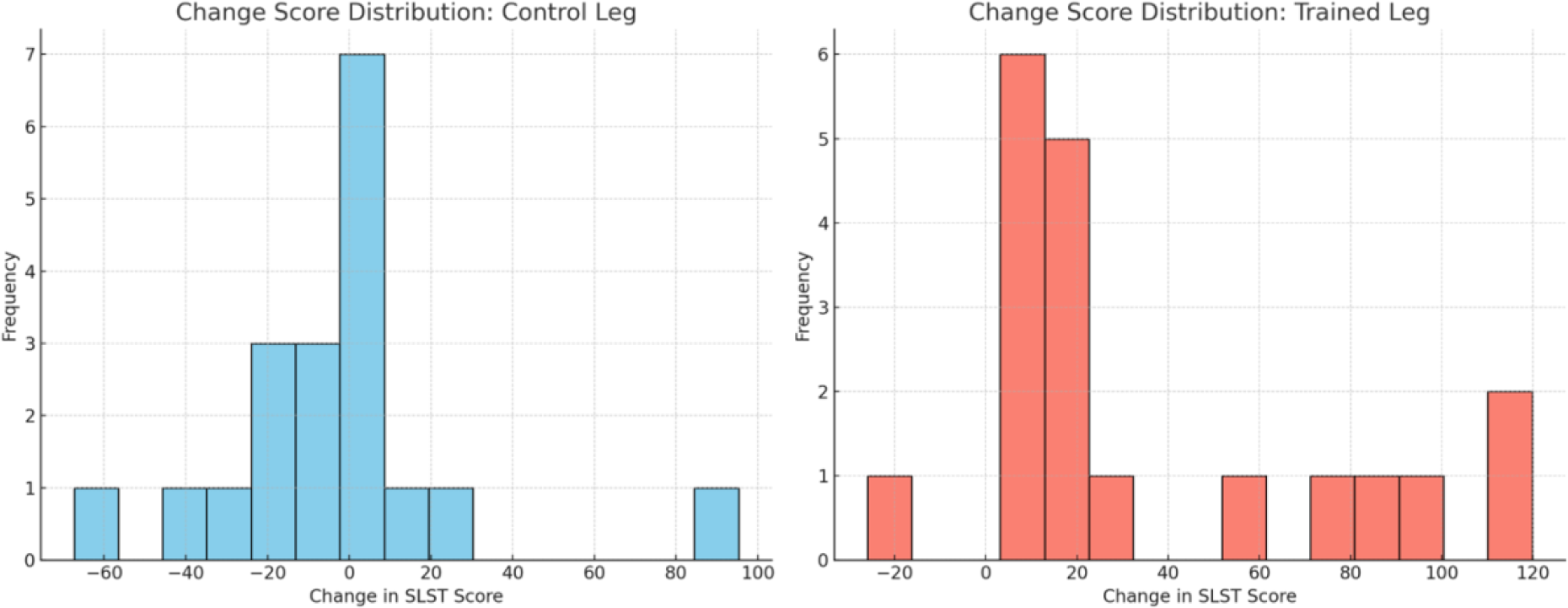
Distribution of SLST Score Changes for Control and Trained Legs. The histograms display the frequency of change in SLST scores from the baseline to the post-training assessment.

Plots of SLST changes and distributions showed that, for the control leg group, most participants displayed little to no change in balance performance (P=1; Sign test). In stark contrast, the trained leg group showed a clear rightward shift, indicative improved balance performance. This contrast highlights the effectiveness of the ankle rotation training in enhancing balance capabilities.

We further conducted multiple linear regression analysis on the potential influences from demographic, physical attributes, and baseline capabilities on training related SLST durations. The regression analysis revealed that pre-training SLST scores, from both the right and left legs, were the most significant predictors of post-training performance. This was a consistent finding across all employed models, indicating the importance of an individual’s baseline balance ability in their potential for improvement.

In addition to the pre-training scores, physical attributes such as limb length and height were also found to be significant factors. These variables underscored the impact of physiological differences among individuals on their response to the balance training.

**Figure 4:**
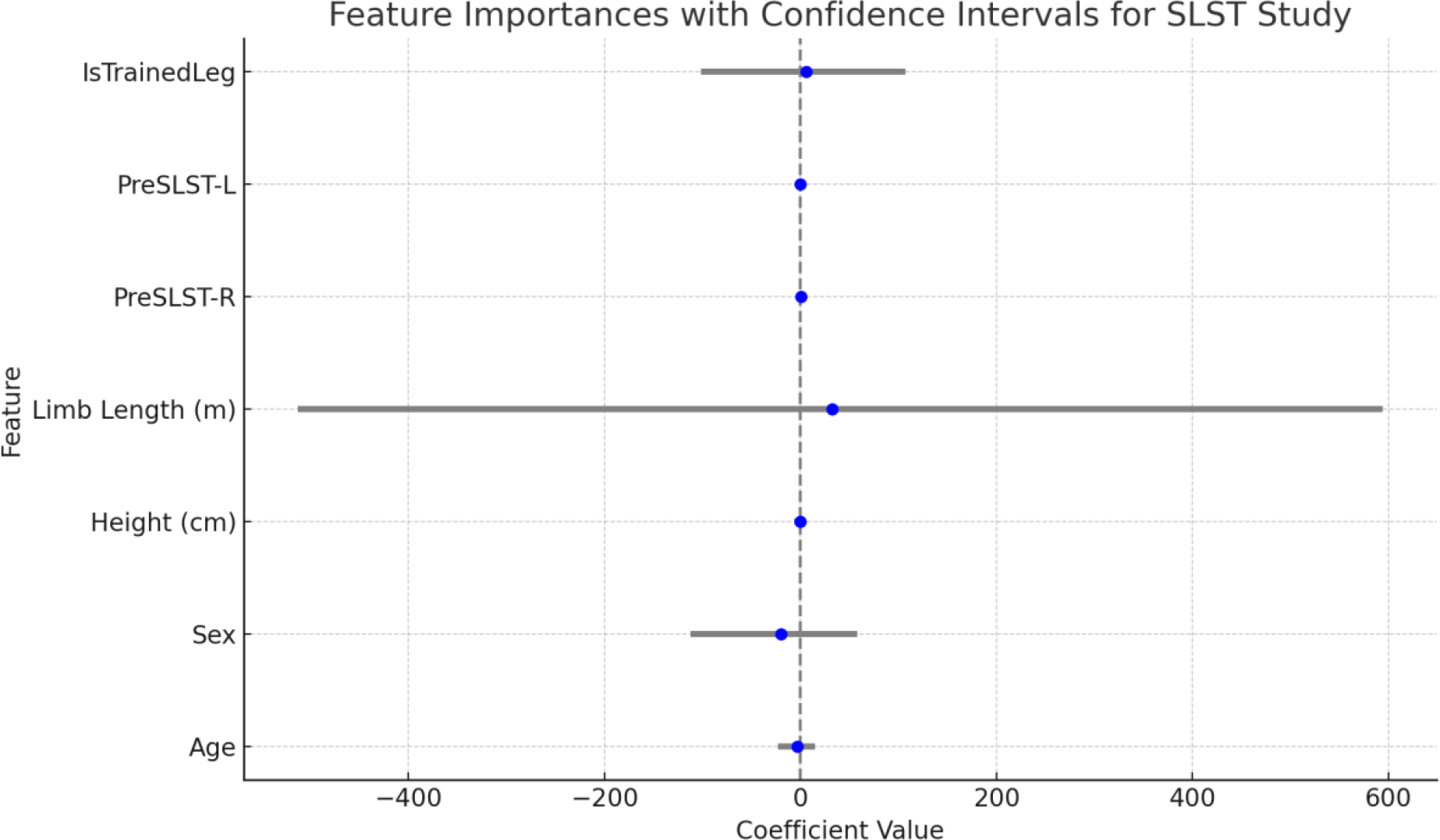
Feature Importances and Confidence Intervals in Predicting SLST Score Changes. This graph illustrates the mean coefficients and 95% confidence intervals for each predictor variable in the SLST study. The coefficients indicate the average impact of each feature on the change in SLST scores, while the horizontal error bars represent the range of the 95% confidence intervals. Features with confidence intervals crossing the zero line suggest less certainty in their predictive power. The plot highlights the complexities in determining the key factors influencing SLST performance improvements, underscoring the multifactorial nature of balance and stability in response to training.

**Figure 5:**
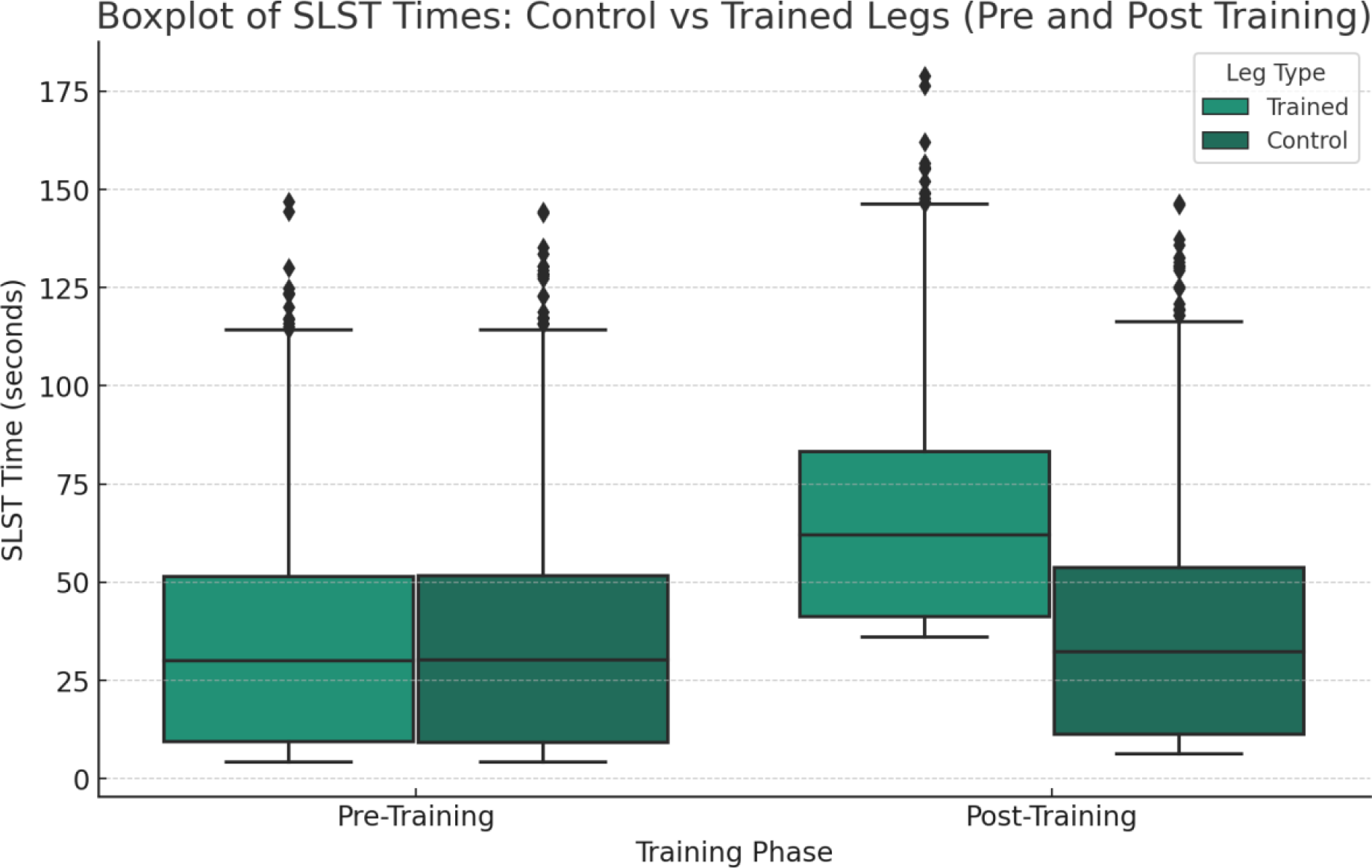
Comparative Boxplot of Single Leg Stand Test (SLST) Times for Trained vs. Control Legs Pre and Post Training. This figure illustrates the distribution of SLST times for both trained and control legs before (Pre-Training) and after (Post-Training) a targeted training intervention. The boxplots provide a visual representation of the central tendency and variability within each group, highlighting median values and interquartile ranges. Notable are the potential outliers, depicted as points beyond the whiskers, which indicate variations in individual performance. The comparison between the ’Trained’ and ’Control’ legs across the two time points offers insights into the training’s impact, with particular attention to shifts in median values and the spread of data post-training.

### Synthetic Data for Large-Scale Analysis

Building upon the pilot data from 18 individuals, we aimed to extrapolate our findings to a broader population. To achieve this, we generated a synthetic dataset that mirrored the statistical properties of our original sample from the Single Leg Stand Test (SLST) intervention study. This approach enabled us to conduct large-scale analyses and draw more generalizable conclusions.

The synthetic dataset creation followed a rigorous process, ensuring that the generated data closely resembled the demographic and SLST characteristics of the original study participants.

Demographic Distribution: The age and sex distributions in the synthetic dataset were modeled to reflect the original data. Ages were generated using a normal distribution centered around the mean age of the original participants, constrained by the observed age range. The sex ratio was maintained as per the original study’s distribution.

Baseline SLST Times: For SLST times, we applied normal distributions with means and standard deviations derived from the original data. The generation process respected the study’s design by excluding any synthetic SLST times below the 2-second minimum.

Training Effect Changes: The training effects on SLST times for both the control and trained legs were synthesized to mirror the mean values recorded in the pilot data, ensuring the synthetic dataset reflected the true impact of the intervention.

Ratios Between Legs’ Times: Ratios of pre- and post-training times between the right and left legs were calculated to ensure the synthetic data did not contain unrealistic differences between the legs, hence preserving the integrity of the original data’s findings.

Dataset Validation: A thorough validation process compared the synthetic dataset against the original data across demographic distributions, baseline SLST times, and training effect sizes. Adjustments were made to the synthetic SLST times to better match the original data and to minimize the occurrence of unrealistically low times.

Final Dataset Composition: The final synthetic dataset consists of 5000 records, mirroring the original dataset’s structure, including demographic information, baseline and post-training SLST times, while ensuring anonymity through the use of codenames.

Ethical Considerations: Throughout the synthetic data generation process, ethical guidelines were rigorously followed to ensure no real participant data was disclosed. The dataset is designed solely for research purposes, offering a valuable asset for simulations and explorations of SLST outcomes, circumventing the ethical and logistical constraints associated with traditional clinical trials.

Synthetic Data Generation Process: starting from the demographic distribution, the process involves modeling age and maintaining sex ratio, generating baseline SLST times with normal distributions and excluding times below 2 seconds, and applying training effects that mirror the original data’s mean values. Ratios between right and left legs’ SLST times ensure realistic bilateral balance, followed by dataset validation to align with the original data, including adjustments of synthetic SLST times. The final dataset composition includes 5000 records that maintain anonymity and adhere to ethical guidelines, facilitating extensive research applications while preserving participant confidentiality.

### Synthetic Data Analysis

#### Synthetic Data Descriptive Statistics

The synthetic data generated for this study presents a detailed overview of various demographic and performance characteristics associated with the Single Leg Stand Test (SLST). This dataset includes information on 5000 synthetic individuals and offers valuable insights into the impact of targeted ankle joint rotation training on balance.

In terms of age and sex distribution, the mean age of participants is 19.34 years, with a standard deviation of 2.24 years. This indicates a moderate age variability within the group, with ages ranging from 15 to 24 years, covering the late adolescence to early adulthood period. Regarding gender, there are 2631 female participants, constituting approximately 52.62% of the sample, and 2369 male participants, making up about 47.38%. This distribution ensures a balanced representation of both genders, which is crucial for generalizing the findings of the study. The Single Leg Stand Test (SLST) times, both before and after the training, are key indicators in this study. Initially, the mean SLST time for the right leg was 35.52 seconds, with a standard deviation of 27.44 seconds, suggesting a wide range of balance times. The left leg had a mean of 33.27 seconds and a standard deviation of 23.55 seconds, showing slightly less variability than the right leg.

Post-training, there was a noticeable improvement in SLST times. The mean time for the right leg increased to 52.61 seconds, with a standard deviation of 31.22 seconds, indicating an increased average balance time post-training. Similarly, the left leg showed improvement, with a mean time of 50.29 seconds and a standard deviation of 27.78 seconds post-training.

These SLST times, both pre- and post-training, provide essential insights into the effectiveness of ankle joint rotation training. The increase in mean SLST times post-training for both legs underscores the potential impact of such exercises in enhancing balance.

### Synthetic Data Analysis

In the examination of the synthetic dataset, which expanded the sample size to 5000 individuals, statistical tests were employed to analyze the Single Leg Stand Test (SLST) performance both before and after the training intervention.

In the analysis of our synthetic dataset, initially modeled to reflect the statistical properties and normal distribution of SLST times as observed in the original study, we encountered a significant challenge due to the presence of extreme values. These outliers, likely picked up while modeling the original data, while aligning with the real-world variability often seen in human performance data, deviated substantially from the normal distribution. This deviation necessitated a reevaluation of our statistical approach, leading us to opt for the Wilcoxon signed-rank test. This non-parametric method is particularly suited for such scenarios as it offers robustness against outliers and is appropriate for analyzing paired observations, like our pre- and post-training SLST times. By employing this test, we ensured a more accurate and reliable analysis that accommodates the full spectrum of data variability, including the extreme values, thereby enhancing the overall validity and relevance of our study’s findings.

A comparative study of pre-training SLST times between the trained and control legs using the Wilcoxon Signed-Rank Test showed no significant difference (p = 0.8317), suggesting that the two groups were comparable at baseline. In contrast, post-training data exhibited a significant improvement in SLST times for the trained legs when compared to the control legs (p < 0.001, Wilcoxon Test). This improvement highlights the effectiveness of the ankle training intervention across the synthetic dataset.

To quantify the size of the training effect, Cliff’s Delta was calculated, revealing a negligible effect size before training (Delta = -0.0005), which aligns with the Wilcoxon Test results indicating no initial differences. Notably, the effect size after training was found to be medium to large (Delta = 0.587), signifying a substantial impact of the training on SLST performance. These results from the synthetic data align with the findings from the original 18-participant dataset, providing further evidence that targeted ankle training can significantly enhance balance as measured by SLST, with the effect size being more pronounced after the training intervention.

### Synthetic Database and Simulated Trials

#### Context and Objective

This section of the paper presents the results of a simulated multi-center clinical trial designed to examine the impact of sample size on statistical conclusions in clinical research. The trial comprised 10 nominal sites, divided into 5 small sample sites (each with 10-30 subjects) and 5 large sample sites (each with a sample size 10 times larger than its corresponding small sample site). The subjects were randomly selected from a synthetic database containing Single Leg Stand Test (SLST) data. The aim was to demonstrate how larger sample sizes might affect statistical outcomes and the influence of covariants.

#### Methodology

Subjects at each trial site underwent the same SLST protocol, and their data were analyzed to assess changes in SLST performance post-training. The analysis included descriptive statistics and paired t-tests. Covariates such as age and sex were also considered in the analysis to evaluate their influence on the outcomes.

## Results

### Small Sample Site Analysis (Sample Range: 10-30 Subjects) Site 1 Example: (20 Subjects)

**Table 2.** Comparative Analysis of Single Leg Stand Test (SLST) Times for a Small Sample Site with 20 Subjects. This table presents the mean SLST times before and after training for both the trained leg and the control leg. Statistical significance is indicated by the T-statistics and P-values, highlighting the impact of the training regimen on the trained leg compared to the untrained control leg. Similar trends were observed in other small sample sites with varying degrees of statistical significance, generally demonstrating improvement post-training but with wider confidence intervals and less robust P-values.

| Leg | Pre-training Mean SLST Time | Post-training Mean SLST Time | T-Statistic | P-Value |
| --- | --- | --- | --- | --- |
| Trained Leg | 43.32 seconds | 63.61 seconds | -6.55 | 1.39e-06 |
| Control Leg | 32.13 seconds | 45.95 seconds | -4.46 | 0.0002 |

### Large Sample Site Analysis (Sample Range: 100-300 Subjects) Site 1 Example: (200 Subjects)

**Table 3:** Comparative Analysis of Single Leg Stand Test (SLST) Times for a Large Sample Site with 200 Subjects. This table outlines the mean SLST times before and after training for both the trained right leg and the control left leg in a large sample size. The T-statistics and P-values demonstrate significant changes post-training, reflecting the effects of the training on the larger cohort.

| Leg | Pre-training Mean SLST Time | Post-training Mean SLST Time | T-Statistic | P-Value |
| --- | --- | --- | --- | --- |
| Trained Leg | 43.32 seconds | 63.61 seconds | -6.55 | 1.39e-06 |
| Control Leg | 32.13 seconds | 45.95 seconds | -4.46 | 0.0002 |
| Right Leg (Trained) | 34.40 seconds | 49.38 seconds | -15.37 | 4.12e-37 |
| Left Leg (Control) | 33.11 seconds | 52.24 seconds | -19.62 | 5.90e-51 |

The large sample sites consistently showed more statistically significant results with tighter confidence intervals. The increased sample size provided more power to detect changes, leading to more reliable statistical conclusions. Furthermore, as shown in Figure 8, there is a strong correlation between baseline SLST durations and training-induced improvements.

**Figure 6:**
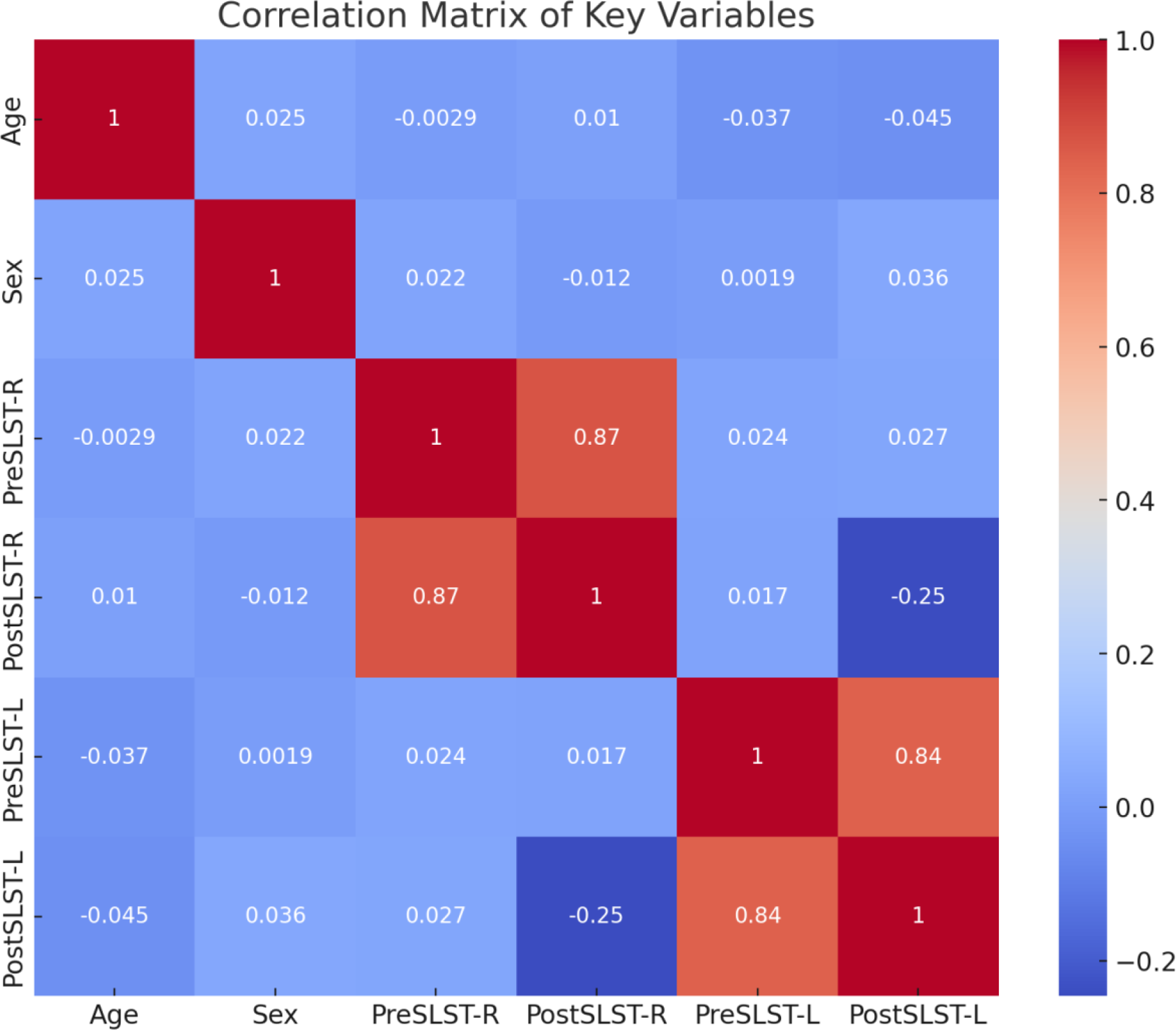
Correlation Matrix of Key Variables in Synthetic Data Analysis. This heatmap provides a visual representation of the correlation coefficients between age, sex, and Single Leg Stand Test (SLST) scores before (PreSLST) and after (PostSLST) the training intervention for both right (R) and left (L) legs. The color gradient from red to blue represents the range from positive (1.0) to negative (-1.0) correlations. Strong positive correlations are evident between the pre- and post-training SLST scores for each leg, whereas age and sex show minimal correlation with SLST scores, indicating that the SLST improvements are predominantly related to the training rather than demographic factors

**Figure 8:**
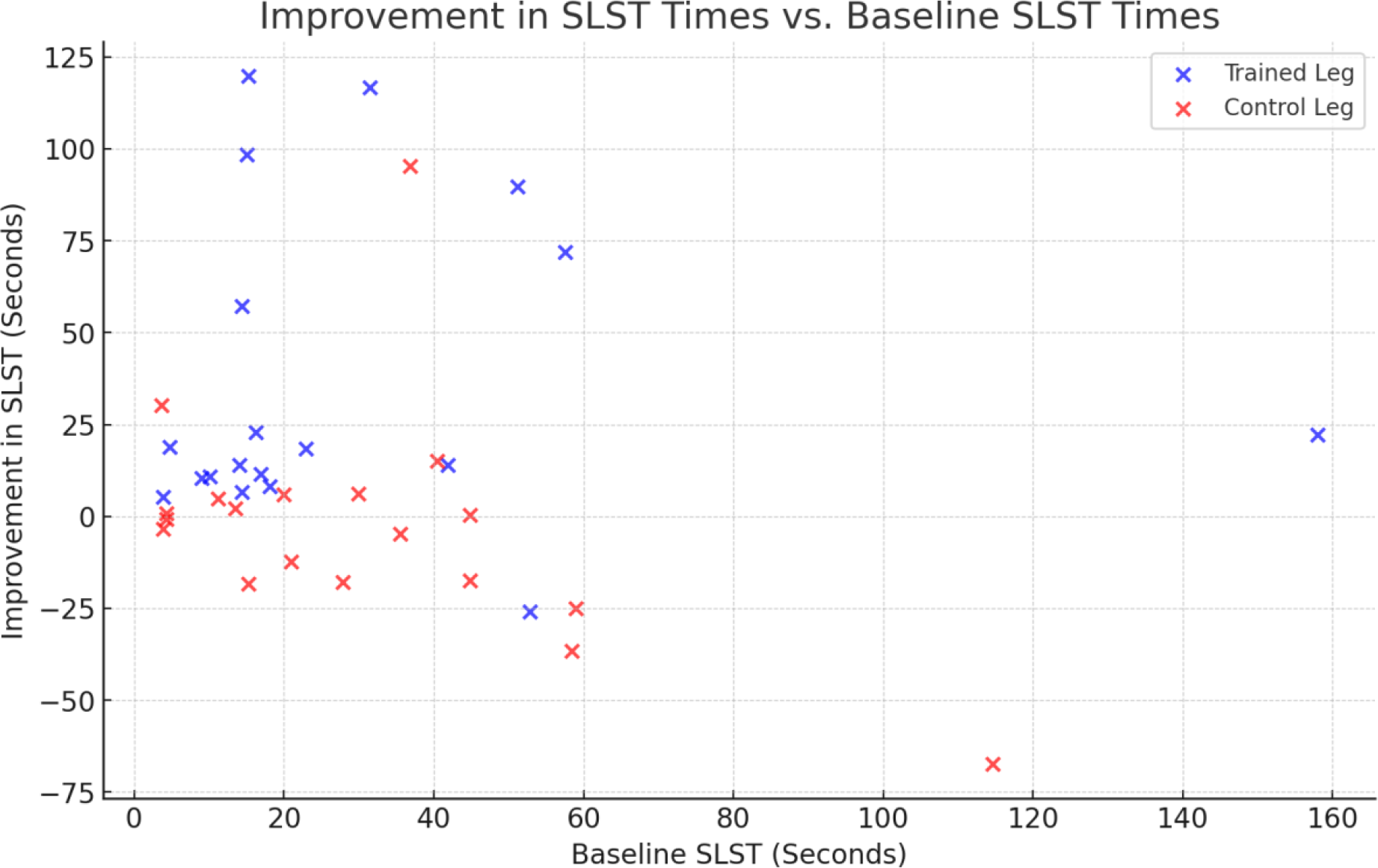
Correlation Between Baseline SLST Durations and Training-Induced Improvements. This scatter plot displays each participant’s baseline SLST duration against the magnitude of improvement post-ankle joint rotation training. A visible trend suggests that individuals with specific baseline SLST durations are more likely to experience substantial improvements, emphasizing the predictive value of initial balance assessments. The differential markers represent the trained and control limbs, illustrating the targeted training’s impact compared to the untrained state.

### Comparative Analysis

Statistical Power and Significance: Larger sample sites demonstrated greater statistical power, evidenced by more significant T-statistics and smaller P-values. This indicates a higher likelihood of detecting true effects of the training protocol.

Effect Size and Confidence Intervals: The larger sample sizes also resulted in more precise estimates of effect sizes and narrower confidence intervals, suggesting more accurate and reliable conclusions.

Covariate Influence: Analysis of covariants like age and sex showed that their influence on SLST performance was more precisely quantified in larger sample sites. This precision is crucial for understanding and controlling for potential confounding factors in clinical research.

## Discussion

The findings from this study demonstrate that targeted ankle single joint rotation training, facilitated by the innovative Ambulosono sensor, significantly enhances balance capabilities, as reflected in the prolonged durations of the Single Leg Stance Test (SLST) in the trained limbs. The Ambulosono sensor’s role has been pivotal in not only facilitating this targeted training but also in ensuring rigorous adherence and engagement, a factor that is particularly critical when considering the essential role of ankle dorsiflexion and plantarflexion in daily activities and fall prevention.

Our findings are consistent with the consensus that improvements in ankle range of motion (ROM) are intrinsically linked to enhanced balance and stability.24 The significant effect size observed for the trained limbs is of both statistical and clinical importance, in stark contrast to the moderate and non-significant changes in the control limbs, thereby underscoring the effectiveness of the intervention. Interestingly, the initial SLST scores have emerged as strong predictors of post-training outcomes, suggesting that intrinsic balance abilities could play a decisive role in the training’s effectiveness. While the performance consistency in control limbs served as a robust internal benchmark, the substantial improvements in the trained limbs point unequivocally to the benefits of ankle joint rotation training. Addressing a gap in existing research, our study’s findings suggest that even with a flexible training schedule, participants can achieve notable balance improvements.

The study’s findings, demonstrating significant improvements in Single Leg Stance Test (SLST) durations, suggest that home-based rehabilitation programs can be as effective as those conducted in clinical environments. This aligns with a growing body of literature, including studies such as that by Lopez-Lira et al., which indicate that at-home rehabilitation for conditions like total knee replacement can achieve results comparable to in-clinic interventions.25, 26 The implications of at-home rehabilitation are profound, particularly considering the possible alleviation of the healthcare system’s burden. By enabling patients to perform rehabilitation exercises in their own homes, there is a reduction in the necessity for physical resources and personnel in clinical settings. This shift not only frees up valuable healthcare resources but also reduces the economic and logistical strain on both patients and healthcare providers.

## Synthetic Database

A pivotal element in enhancing the robustness of this study’s findings is the creation and utilization of a synthetic database, created and validated using the original data, which greatly facilitated the research process. By generating a synthetic dataset of 5000 individuals, the study significantly increased its sample size, moving beyond the limitations of the original smaller dataset. This expansion is crucial for achieving a more accurate and realistic estimation of effect sizes and for capturing a wider range of variability in human balance and mobility responses. The synthetic database allows for a more comprehensive exploration of potential outcomes and relationships within the data, providing a deeper and more nuanced understanding of the impact of ankle joint rotation training on balance. This approach not only enriches the study’s statistical power but also broadens its applicability and relevance to a larger population, making the findings more generalizable and robust. Employing a synthetic database thus represents a strategic advancement in research methodology, offering a scalable and ethically sound approach to simulate extensive and diverse datasets for advanced scientific analysis.

Lastly, GPT’s ability to analyze and interpret high-dimensional data can demystify complex concepts, making statistics more approachable for students. By visualizing data, identifying patterns, and summarizing findings in a way that is easy to understand, GPT can help bridge the gap between abstract statistical theories and their practical applications.

In conclusion, the integration of GPT into undergraduate statistics and research training offers a wealth of benefits, from personalized learning experiences to exposure to advanced analytical tools. This innovative approach not only enriches the educational journey of students but also equips them with the skills necessary to thrive in the data-driven landscape of the future.

## Limitations and Considerations

This study’s insights are contextual and may not be universally applicable to people of an older age or patients with balance impairments. The study also suffered from the limitations of the dataset size and the specificity of the exercise modality studied. Further research is not only needed to validate the effectiveness of ankle training paradigm but confirm that enhanced SLST can translate into a reduction of fall and injury during daily living.

Finally, future investigations into the Ambulosono sensor’s specific contributions are also needed to better understand technology’s potential in enhancing at-home rehabilitation outcomes. The exploration into sensor-based compliance supports could inform the development of more effective, patient-centric rehabilitation protocols that capitalize on the convenience and accessibility of at-home care while maintaining the rigor and effectiveness of traditional clinical rehabilitation methods.

## Data Availability

All data produced in the present study are available upon reasonable request to the authors

## Acknowledgments

This research was partially supported by the grants from Alberta Ministry of Mental Health and Hotchkiss Brain Institute of the University of Calgary.

## References

[1] Morgan K. The use of a tri-axial accelerometer to measure changes in lower extremity fatigue during functional activity. Theses and Dissertations [Internet]. 2009 Aug 5 [cited 2023 Sep 18]; Available from: https://scholarscompass.vcu.edu/etd/1924/

[2] Li J, Wei Y, Wei M. Finite Element Analysis of the Effect of Talar Osteochondral Defects of Different Depths on Ankle Joint Stability. Medical Science Monitor. 2020 Jun 20;26.

[3] Deschamps K, Karel Mercken, Verschuren PM, Maarten Eerdekens, Eline Vanstraelen, Sander Wuite, et al. Foot biomechanics in patients with advanced subtalar- and mid-tarsal joint osteoarthritis and poorly responding to conservative treatment. Journal of Foot and Ankle Research. 2023 Nov 28;16(1).

[4] Gatti R, Corti M, Cervi P, Pulici L, Boccardi S. Biomechanics of lower limb raising from the supine position. Europa Medicophysica [Internet]. 2006 Sep 1 [cited 2023 Dec 6];42(3):185–93. Available from: https://pubmed.ncbi.nlm.nih.gov/17039214/

[5] Lawford BJ, Dobson F, Bennell KL, Merolli M, Graham B, Haber T, et al. Clinician-administered performance-based tests via telehealth in people with chronic lower limb musculoskeletal disorders: Test–retest reliability and agreement with in-person assessment. Journal of Telemedicine and Telecare. 2022 Nov 30;1357633X2211373–3.

[6] Cabell L, Pienkowski D, Shapiro R, Janura M. Effect of Age and Activity Level on Lower Extremity Gait Dynamics. Journal of Strength and Conditioning Research. 2013 Jun;27(6):1503–10.

[7] Sadeghi H, Jehu DA, Daneshjoo A, Shakoor E, Razeghi M, Amani A, et al. Effects of 8 Weeks of Balance Training, Virtual Reality Training, and Combined Exercise on Lower Limb Muscle Strength, Balance, and Functional Mobility Among Older Men: A Randomized Controlled Trial. Sports Health: A Multidisciplinary Approach. 2021 Feb 13;194173812098680.

[9] Rubenstein LZ, Josephson KR, Robbins AS. Falls in the nursing home. Annals of internal medicine [Internet]. 1994 [cited 2019 Jun 26];121(6):442–51. Available from: https://www.ncbi.nlm.nih.gov/pubmed/8053619

[9] Tu R, Wang S, He H, Ding J, Zeng Q, Guo L, et al. Association between subjective cognitive complaints, balance impairment and disability among middle-aged and older adults: Evidence from a population-based cohort study. Geriatrics & Gerontology International. 2022 Nov 3;22(12):1025–31.

[10] Montero-Odasso M, van der Velde N, Martin FC, Petrovic M, Tan MP, Ryg J, et al. World guidelines for falls prevention and management for older adults: A global initiative. Age and Ageing [Internet]. 2022 Sep;51(9). Available from: https://academic.oup.com/ageing/article/51/9/afac205/6730755

[11] Matson T, Schinkel-Ivy A. How does balance during functional tasks change across older adulthood? Gait & Posture. 2020 Jan;75:34–9.

[12] Bretan O, Elias Silva J, Ribeiro OR, Corrente JE. Risk of falling among elderly persons living in the community: assessment by the Timed up and go test. Brazilian Journal of Otorhinolaryngology. 2013 Jan;79(1):18–21.

[13] Blodgett JM, Hardy R, Davis D, Peeters G, Hamer M, Kuh D, et al. Prognostic accuracy of the one-legged balance test in predicting falls: 15-years of midlife follow-up in a British birth cohort study. Frontiers in sports and active living. 2023 Jan 9;4.

[14] Cho I, Kaplanidou K, Sato S. Gamified Wearable Fitness Tracker for Physical Activity: A Comprehensive Literature Review. Sustainability. 2021 Jun 22;13(13):7017.

[15] Southard V. A Randomized Control Trial of the Application of Efficacy Training to Balance Assessment. Physical & Occupational Therapy In Geriatrics. 2006 Jan;25(2):51–66.

[16] McColl L, McMeekin P, Poole M, Parry SW. Is fear of falling key to identifying gait and balance abnormalities in community-dwelling older adults? Protocol of a mixed-methods approach. BMJ Open. 2022 Dec;12(12):e067040.

[17] Stevens JA, Corso PS, Finkelstein EA, Miller TR. The costs of fatal and non-fatal falls among older adults. Injury Prevention [Internet]. 2006 Oct 1 [cited 2019 Jul 21];12(5):290–5. Available from: https://www.ncbi.nlm.nih.gov/pmc/articles/PMC2563445/

[18] Krueger B, Becker L, Leemkuil G, Durall C. Does Talocrural Joint-Thrust Manipulation Improve Outcomes After Inversion Ankle Sprain? Journal of Sport Rehabilitation. 2015 Aug;24(3):315–21.

[19] Blodgett JM, Hardy R, Davis D, Peeters G, Hamer M, Kuh D, et al. Prognostic accuracy of the one-legged balance test in predicting falls: 15-years of midlife follow-up in a British birth cohort study. Frontiers in sports and active living. 2023 Jan 9;4.

[20] Hertel J. Functional Anatomy, Pathomechanics, and Pathophysiology of Lateral Ankle Instability. Journal of athletic training [Internet]. 2002;37(4):364–75. Available from: https://www.ncbi.nlm.nih.gov/pmc/articles/PMC164367/

[21] Kasli K, Devrim Sahin C, Ilcin N. The Short-Term Effect of Myofascial Relaxation by Roller Massage on Ankle Joint Range of Motion in Older Adults. Topics in Geriatric Rehabilitation. 2022 Jan;38(1):35–41. Muhammad Farhan Raza (30175374)

[22] Willwacher S, Bruder A, Robbin J, Kruppa J, Mai P. A Multidimensional Assessment of a Novel Adaptive Versus Traditional Passive Ankle Sprain Protection Systems. The American Journal of Sports Medicine. 2023 Feb 3;51(3):715–22.

[23] Chomiak T, Sidhu A, Watts A, Su L, Graham B, Wu J, et al. Development and Validation of Ambulosono: A Wearable Sensor for Bio-Feedback Rehabilitation Training. Sensors. 2019 Feb 8;19(3):686.

[24] Wang J, Zhang D, Zhao T, Ma J, Jin S. Effectiveness of balance training in patients with chronic ankle instability: protocol for a systematic review and meta-analysis. BMJ Open. 2021 Sep;11(9):e053755.

[25] Hsieh Y, Chang K, Hung J, Wu C, Fu M, Chen C. Effects of Home-Based Versus Clinic-Based Rehabilitation Combining Mirror Therapy and Task-Specific Training for Patients With Stroke: A Randomized Crossover Trial. Archives of Physical Medicine and Rehabilitation. 2018 Dec;99(12):2399–407.

[26] López-Liria R, Padilla-Góngora D, Catalan-Matamoros D, Rocamora-Pérez P, Pérez-de la Cruz S, Fernández-Sánchez M. Home-Based versus Hospital-Based Rehabilitation Program after Total Knee Replacement. BioMed Research International [Internet]. 2015;2015. Available from: https://www.ncbi.nlm.nih.gov/pmc/articles/PMC4415465/

